# Identification of the motivators and barriers to cochlear implantation in adults over 60 years at an auditory implant centre in the UK: a mixed-methods study

**DOI:** 10.64898/2026.01.08.26343668

**Authors:** Alisha Giby, Kate Hough, Callum Findlay, Mary Grasmeder, Tracey Newman

**Author notes:** Corresponding author: Tracey A Newman, Faculty of Medicine, Building 85, Highfield Campus, University of Southampton, UK, SO17 1BJ 02380599665.

## Abstract

**Objectives:** Cochlear implants are an effective intervention for people with severe to profound hearing loss. However, only a small percentage of those who could benefit from cochlear implants have one. We aimed to understand the barriers to access and receipt of cochlear implants in the UK.

**Design:** Mixed-methods study.

**Setting:** The University of Southampton Auditory Implant Service.

**Participants:** Data on referral route and personal characteristics of 456 patients over the age of 60 at the time of cochlear implantation, who received a cochlear implant before 2020 were included. Semi-structured interviews were held with six people who hear with an implant.

**Primary Outcome Measures:** Demographic factors and routes of referral for cochlear implantation for older adults who went on to receive an implant. Semi-structured interviews were designed to identify key motivators and barriers to receiving a cochlear implant.

**Results:** Sex and ethnicity did not affect cochlear implant uptake, whereas socioeconomic status and differences in referral pathways were associated with differences in uptake. People from lower socioeconomic groups were underrepresented in the implanted population at USAIS. Certain health providers across the cohort catchment area were more likely to refer patients than others which in turn affected cochlear implant uptake. Barriers to uptake were poor knowledge about implants by patients and clinicians, and fear of surgery. A willingness by patients to explore a way to reduce the daily challenges associated with hearing loss and the support and encouragement of clinicians, family and friends and other people with implants were motivators to implant uptake.

**Conclusion:** These findings will inform future research to address the key factors preventing eligible individuals from receiving cochlear implants. This will support the development of strategies to improve access to, and uptake of, cochlear implants for older adults.

**ARTICLE SUMMARY:** *Strengths and limitations of this study:*

- Strength: Mixed-methods approach was used:

- A service evaluation where we analysed retrospective data from 456 patients from a UK cochlear implant centre.
- Semi-structured interviews were used to gather rich qualitative data for a deeper insight into the barriers and motivators of cochlear implant uptake.
- Limitation: Data from patients that were referred to USAIS but did not receive cochlear implants was not included.
- Limitation: Sample size for interviews was small (six participants) due to time constrains of the (student) project.

## INTRODUCTION

Hearing loss affects over 1.5 billion people globally (1). There are an estimated 1.2 million people in the UK living with severe or profound hearing loss. Hearing loss is a major cause of years lived with disability and contributes to a reduced quality of life (2). It is estimated that hearing loss will cost the UK economy £38.6 billion due to lost productivity in the year 2031 (3). Cochlear implants (CI) are recommended by the National Institute for Health and Care Excellence (NICE) for adults with severe to profound sensorineural hearing loss who do not receive adequate benefit from hearing aids (4). Cochlear Implants are surgically implanted into the inner ear to replace the function of damaged hair cells in the cochlea. Electrodes on the implant stimulate the auditory nerve which the user perceives as sound. In the UK, cochlear implant assessment and surgeries are performed at 19 specialised cochlear implant centres, one of which is the University of Southampton Auditory Implant Service (USAIS). This service covers the South of England and Channel Islands. It receives CI referrals from NHS and private audiologists, general practitioners (GPs) and Ear, Nose and Throat (ENT) specialists.

Implantation is a cost-effective intervention (5), and the expected benefits are improvements in communication and relationships, access to education and employment, independence in daily life and a reduction in long-term cognitive decline (6–8). Estimates are that around only 5% of eligible adults in the UK, i.e., those who are within clinical criteria and who may benefit, undergo implantation (9). Barriers to access to cochlear implants (10–12), have been identified. Key amongst these in the UK is low rates of referral, with only 9% of clinically eligible deafened adults being referred (13). People less likely to be referred included those living in more deprived areas, males and those from ethnic minorities (13). Studies conducted outside the UK suggest poor public awareness of hearing loss and cochlear implants, patients’ concerns about surgery and practical factors contribute to low uptake of implants (10–12,14). Despite this understanding, there is a gap in cochlear implant research as to the reasons for demographic differences and other factors which might influence patients’ access to, and uptake of, implantation in the UK. This mixed-methods study aims to evaluate the demographics and routes of referral for cochlear implantation for older adults (60 years and above) under the care of the University of Southampton Auditory Implant Service (USAIS) and to explore motivators and barriers that inform older adults’ decision-making about cochlear implantation.

## METHODS

This study was conducted in two phases: a quantitative service evaluation and qualitative semi-structured interviews.

### Service evaluation

A service evaluation of University of Southampton Auditory Implant Service (USAIS) was carried out to investigate the demographic profile of patients and assess the referral pathways through which patients were referred to the service. Ethical approval for the service evaluation was granted by the University of Southampton Ethics committee (ERGO II: 76664).

Data were sourced from USAIS electronic patient records of all patients at USAIS who had received a cochlear implant by 2021. The inclusion criteria included patients aged 60 years or over when referred to USAIS and the exclusion criteria included patients who received their implant at another CI centre. The study size was determined by the number of patients at USAIS that met the inclusion criteria. Supplementary figure 1 details how the inclusion and exclusion criteria were applied to the patient data.

Data was extracted into Microsoft Excel which included: Sex, Ethnicity, Postcode, Date of birth, Date of deafness, Date of implantation, Date of referral, Aetiology of hearing loss, Referral district, Referral region, Referrer’s designation, Referrer’s address. Patient data was pseudonymised.

Postcodes were matched to their corresponding Index of Multiple Deprivation (IMD) decile as published by the Geographic Data Service, as a measure of the patients’ socioeconomic status (15). The IMD denotes the level of deprivation of an area determined using data about income, employment, education, health, crime, barriers to housing and services and the living environment (14). The health deprivation and disability decile, one of the component domains of the IMD, was used as a surrogate indicator of the quality of healthcare services in the area. IMD data was collected from the English Indices of Deprivation 2019 (16) database. IMD data was not available for postcodes in the Channel Islands, so these were not included in socioeconomic analyses.

Descriptive statistics were used to analyse demographic information, socioeconomic status, characteristics of hearing loss, time between deafness and referral, time between referral and implantation, referrer type and referral district. Frequencies and percentages were reported for categorical variables. Mean and standard deviation were calculated for normally distributed continuous variables while median and interquartile range (IQR) were used to describe variables that were not normally distributed.

A Chi-square test of independence was performed to compare sex of the study population to the sex of over 60s in the South of England and the Channel Islands, using data collected from the UK, Guernsey and Jersey census 2021 (17–19). The ethnicities of patients in the study population were compared to the ethnicities of the over 60s population in the South of England using data collected from the UK census 2021 database (20) (this data was not available for the Channel Islands). Fisher’s exact test was performed for the ethnicity analyses as more than 20% of expected cells had values <5. A p-value less than 0.05 was considered statistically significant.

District population data was obtained from the 2020 ONS Clinical Commissioning Group (CCG) population estimates (21). The percentage of referrals received from each district was compared, controlling for the districts’ differing age demographics (21).

Data were analysed in Microsoft Excel version 16.66.1 and SPSS version 28.0.1.1.

For the observational quantitative data from the service evaluation, we used the STROBE reporting guidelines (22) to draft this manuscript, and the STROBE reporting checklist (23) when editing, included as supplementary figure 2.

### Semi-structured interviews

Semi-structured interviews were carried out with patients with implants under the care of USAIS to explore their experiences with hearing loss and their cochlear implant decision-making process. Ethical approval for the semi-structured interviews was granted by the University of Southampton Ethics committee (ERGO II: 80892).

Convenience sampling was used for recruitment to the study. An invitation was shared via email with members of the USAIS Patient and Public Involvement and Engagement (PPIE) group, ALL_EARS@UoS, and advertised in a poster in the main reception at USAIS (supplementary figure 3). Additional snowball sampling was also utilised. The inclusion criteria for the interviews were adults aged 60 or over who had been assessed for a cochlear implant at USAIS and found to be eligible. The exclusion criteria were people not previously under the care of USAIS, those who had been referred to USAIS, but who did not meet eligibility criteria for cochlear implantation and those who were not able to consent to taking part in the study.

Eligible individuals who expressed an interest in the study were sent a Participant Information Sheet and consent form (Supplementary figure 4 and 5) to sign and return if they wanted to participate. Upon receipt of the consent form, participants were offered a face-to-face interview or interview via Microsoft Teams depending on the participant’s preference. Participants were offered a British Sign Language interpreter, stenographer (to provide captions) or language translator to be present during the interview, if required. A confidentiality agreement was signed by the interpreter or translator (supplementary figure 6).

A total of six participants took part in the semi-structured interviews.

A semi-structured interview guide was used as a framework for discussion (See supplementary figure 7). This explored participants’ experiences with hearing loss, receiving a CI and factors that affected their CI decision-making process. Demographic information was also collected. Each interview lasted between 30-45 minutes.

Interviews were audio-recorded and transcribed in Microsoft Word version 16.66.1. The recordings were deleted after being transcribed and the transcripts were assigned a participant number to ensure anonymity. Identifiable names and places were replaced with pseudonyms.

The interview transcripts were uploaded to NVivo 1.7.1 and analysed iteratively using inductive thematic analysis following Braun and Clarke’s framework (24–26). The transcripts were coded line-by-line to develop a coding framework that could be used for subsequent transcripts. From this, codes were collated to identify themes and subthemes to develop the thematic map. Sentences relating to each subtheme were extracted from the transcripts.

### Patient and Public Involvement

ALL_EARS@UoS is a patient and public involvement and engagement (PPIE) group for people with lived experience of hearing loss that was set up by researchers Professor Tracey Newman and Dr Kate Hough at the University of Southampton. The group has been established since March 2022 and now has around 60 active members. Through regular discussions with group members, the diverse challenges that people had faced when accessing cochlear implants became apparent. This research project evolved from discussions around barriers and motivators to cochlear implantation. Group members have been involved across all aspects of the research cycle including the design and planning, recruitment, implementation and dissemination. The research plan was discussed with the group and feedback on written documents were obtained. Group members shared the study information with peers from local support groups to support recruitment. The findings of the study were presented to the group during a meeting in April 2024 and discussed in the context of a multi-centre study (13). Following this, a group member wrote to eight local hospitals to highlight the need for more awareness of cochlear implants and suggested more signposting is needed on their websites. In addition, we published a letter in the BMJ, with ALL_EARS members as co-authors, to outline key deaf awareness strategies that should be implemented in healthcare (27).

## RESULTS

### Service evaluation

#### Demographics analysis

There were 456 people under the care of USAIS over the age of 60 who had received a cochlear implant. Table 1 shows the demographics of patients in the study population. The majority were female (57.7%) and from a White ethnic background (94.5%). The median age of patients at referral to USAIS was 71.79 years (IQR, 65.5-77.6).

**Table 1.** Demographics of the service evaluation study population.

| Demographics | *n* | % |
| --- | --- | --- |
| Sex |  |  |
| Male | 193 | 42.3 |
| Female | 263 | 57.7 |
| Age at referral to USAIS (years) |  |  |
| 60-65 | 122 | 26.8 |
| 66-70 | 87 | 19.1 |
| 71-75 | 94 | 20.6 |
| 76-80 | 81 | 17.8 |
| 81-85 | 46 | 10.1 |
| >86 | 18 | 4.0 |
| Unknown | 8 | 1.8 |
| Ethnicity |  |  |
| British (White) | 374 | 82.0 |
| Irish (White) | 4 | 0.9 |
| Any other White background | 53 | 11.6 |
| Indian (Asian/British Asian) | 4 | 0.9 |
| Caribbean (Black/Black British) | 2 | 0.4 |
| White and Asian (Mixed) | 1 | 0.2 |
| Unknown | 18 | 4.0 |

N=456. Median age at referral to USAIS was 71.79 years (IQR, 65.5-77.6)

When the sex of the study population was compared to the sex of over 60s in the USAIS catchment area, the Chi-square independence test showed no significant difference in sex between these two groups (χ2=3.131; df=1; p=0.077) (Figure 1a). Likewise, when the ethnicity of the study population was compared to the ethnicities of over 60s in the catchment area, there was no significant difference in ethnicities between the two groups (p=0.224) using Fisher’s exact test (Figure 1b).

**Figure 1.**
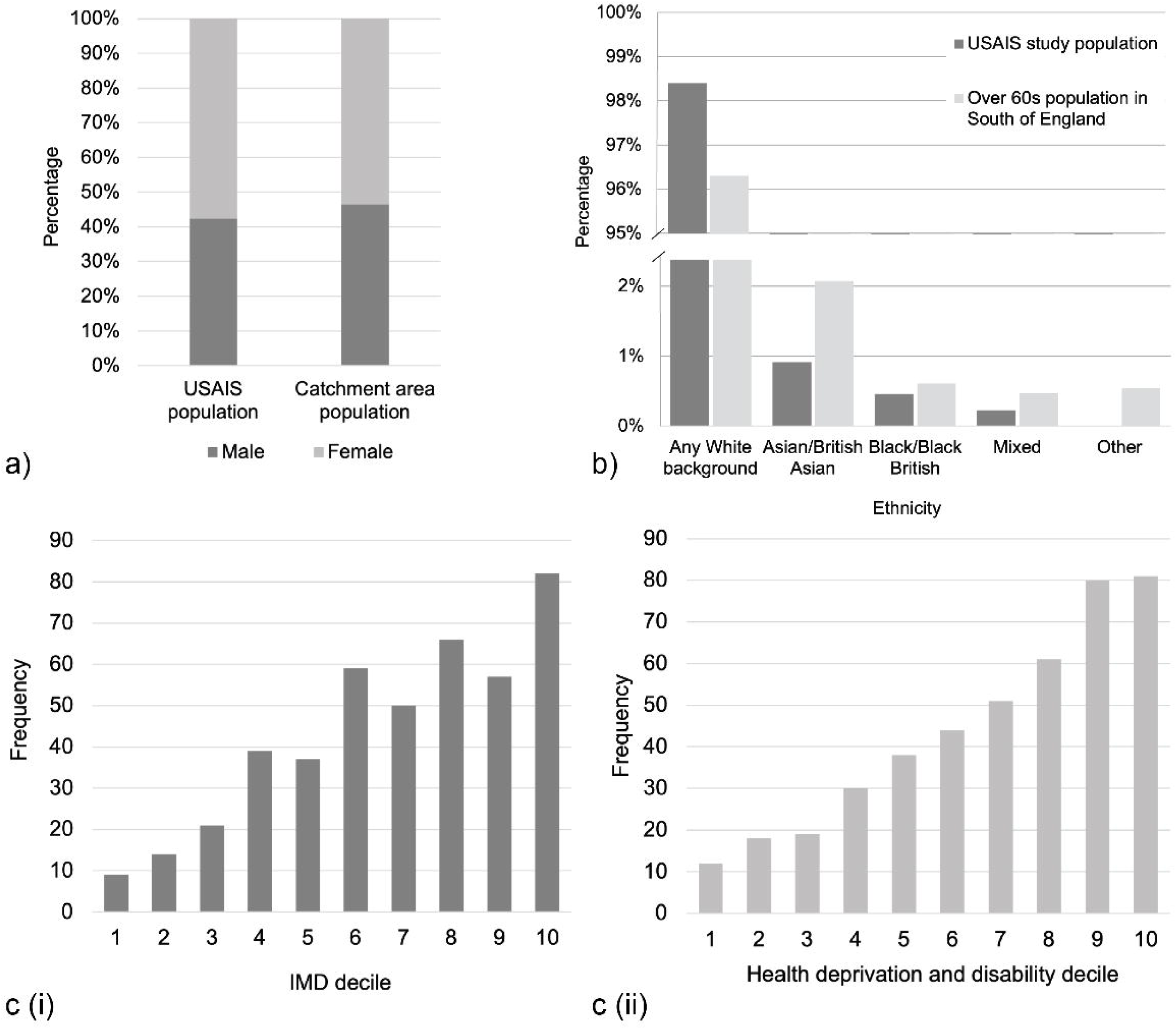
Demographics of the USAIS study population. a. Sex of the USAIS study population (n=456), (B) Sex of over 60s in the South of England and Channel Islands (n=4,024,402). The Chi-square independence test showed no significant difference in proportions of males and females between the groups (χ2 = 3.131; df = 1; p = 0.077). b. Ethnicities of the USAIS study population (dark grey, n=438) compared to ethnicities of over 60s in the South of England (light grey, n=3,980,595). Fisher’s exact test showed no significant difference in proportions of ethnicities between these groups (p= 0.224). c. (i) Number of patients in the USAIS study population in each Index of Multiple Deprivation (IMD) decile (n=434). (ii) Number of patients in the USAIS study population in each health deprivation and disability decile (n=434). 1=most deprived group, 10=least deprived group

On analysis of socioeconomic status, the mean IMD decile of the study population was 7 (IQR, 5-9) and the median health deprivation and disability decile was 8 (IQR, 5-9). The distributions of both variables were negatively skewed, with more patients in the less deprived groups: 47.24% and 51.15% of patients were in deciles 8, 9 or 10 for IMD and health deprivation and disability respectively (Figures 1c(i) and 1c(ii)).

#### Referral pathway analysis

The median time between being diagnosed as deaf and being referred to USAIS was 19.63 years (IQR, 5.42-40.75 years). The median time between referral and implantation was 7 months (IQR, 6-10 months). The percentage of referrals sent to USAIS by each referrer designation (audiologist, ENT specialist or GP) is shown in Figure 2a. Most referrals (66%) were sent by audiologists while only 3% were sent by GPs.

**Figure 2.**
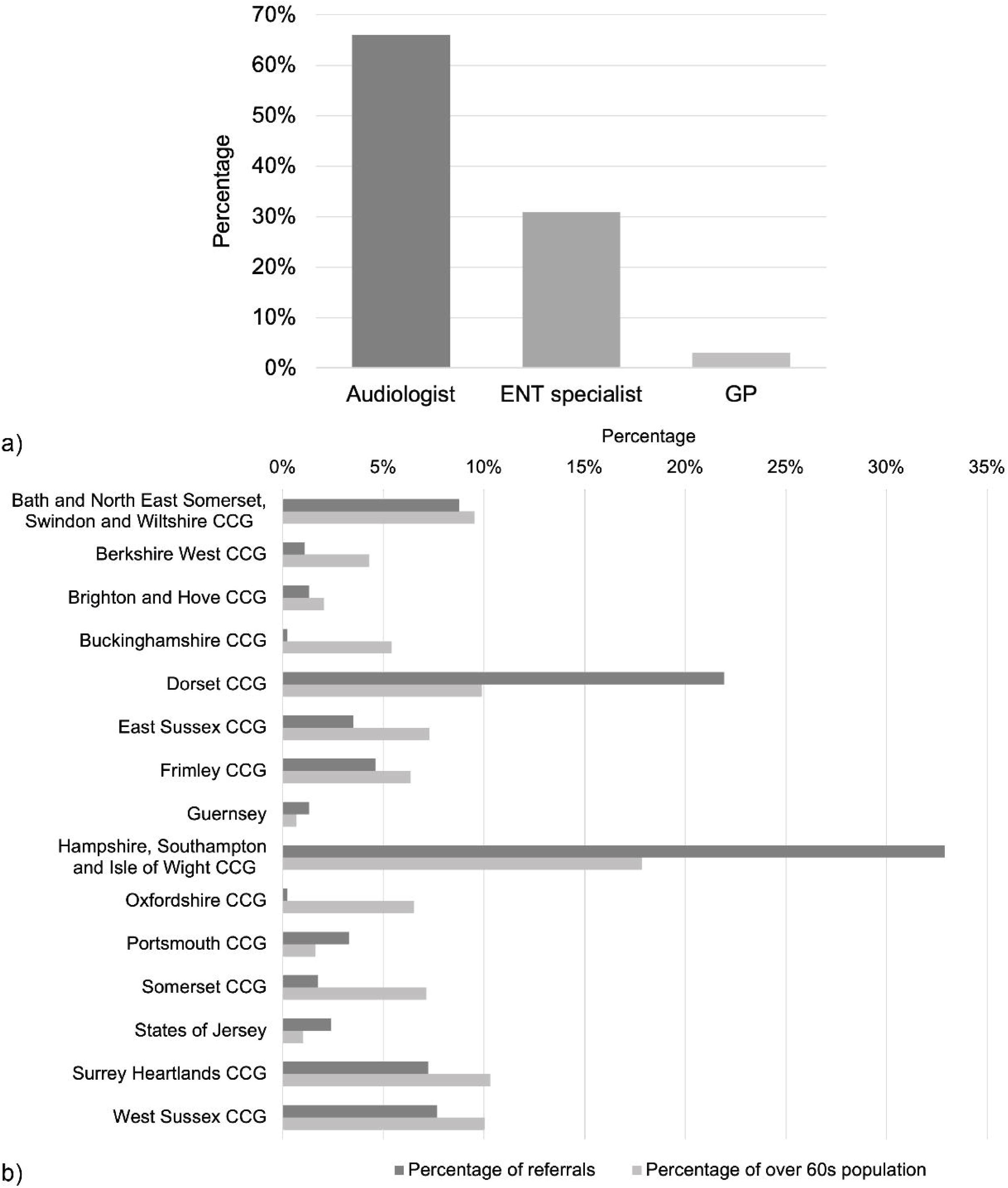
Referrals to USAIS by clinician type and district. a. Distribution by referrer designation (n=262). N.B.: This data was not available for 194 patients. b. Percentage of referrals sent to USAIS by each district* (dark grey, n=456) compared to the percentage of the over 60s population that each district comprises out of the total over 60s population of the catchment area (light grey, n=2,524,566). *Private and unspecified referrals not included in the analysis.

Table 2 shows the number of referrals sent to USAIS by each district or CCG in the South of England and Channel Islands. Figure 2b compares the percentage of referrals that were sent by each district or CCG to the percentage of CCG/district population who are over 60. In some cases, this was disproportionate. For example, Dorset CCG referred 21.93% of patients in the study population but Dorset only comprises 9.88% of the catchment area population of over 60s. On the other hand, Oxfordshire comprises 7.16% of the population but only sent 0.22% of referrals.

**Table 2.**
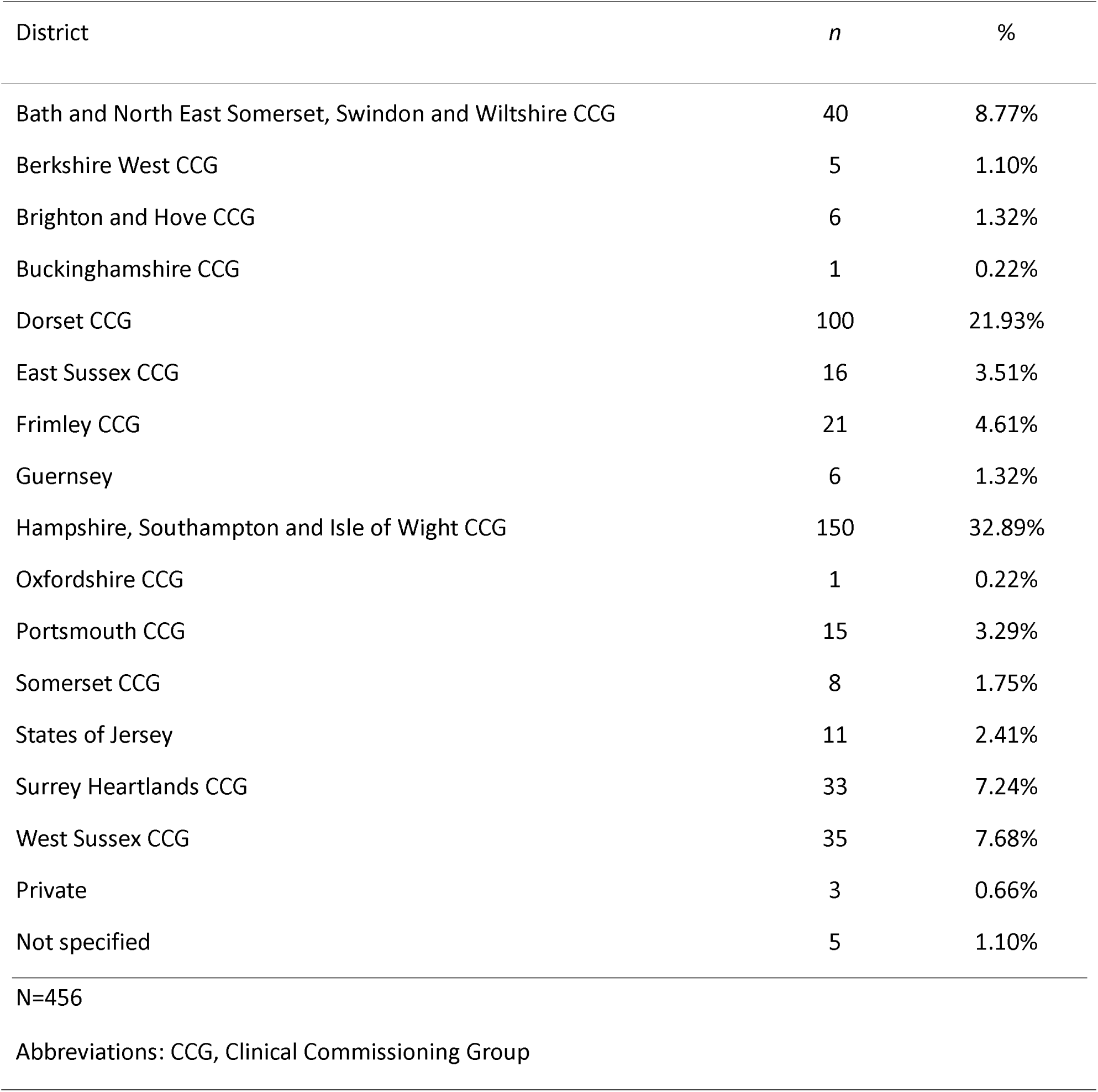
Number of referrals sent by each district.

| District | *n* | % |
| --- | --- | --- |
| Bath and North East Somerset, Swindon and Wiltshire CCG | 40 | 8.77% |
| Berkshire West CCG | 5 | 1.10% |
| Brighton and Hove CCG | 6 | 1.32% |
| Buckinghamshire CCG | 1 | 0.22% |
| Dorset CCG | 100 | 21.93% |
| East Sussex CCG | 16 | 3.51% |
| Frimley CCG | 21 | 4.61% |
| Guernsey | 6 | 1.32% |
| Hampshire, Southampton and Isle of Wight CCG | 150 | 32.89% |
| Oxfordshire CCG | 1 | 0.22% |
| Portsmouth CCG | 15 | 3.29% |
| Somerset CCG | 8 | 1.75% |
| States of Jersey | 11 | 2.41% |
| Surrey Heartlands CCG | 33 | 7.24% |
| West Sussex CCG | 35 | 7.68% |
| Private | 3 | 0.66% |
| Not specified | 5 | 1.10% |
| N=456 |  |  |
| Abbreviations: CCG, Clinical Commissioning Group |  |  |

### Semi-structured interviews

Six cochlear implant (CI) users participated in semi-structured interviews exploring their experiences with hearing loss and the process of being referred, assessed and receiving a CI. The demographics of the participants are shown in Table 3.

**Table 3.** Demographics of interview participants.

| Demographics | *n* | % |
| --- | --- | --- |
| Sex |  |  |
| Male | 4 | 66.67 |
| Female | 2 | 33.33 |
| Age |  |  |
| 60-69 | 2 | 33.33 |
| 70-79 | 3 | 50 |
| 80-89 | 1 | 16.67 |
| >90 | 0 | 0 |
| Ethnicity |  |  |
| White (British) | 6 | 100 |
| Any other White background | 0 | 0 |
| Asian/British Asian | 0 | 0 |
| Black/Black British | 0 | 0 |
| Mixed | 0 | 0 |
| Other | 0 | 0 |
| CI status |  |  |
| CI user | 6 | 100 |
| Undergoing CI assessment | 0 | 0 |
| Declined CI | 0 | 0 |
| N=6 |  |  |

**Table 4.** Themes and subthemes developed from qualitative analysis of interviews.

| Theme | Subtheme |
| --- | --- |
| **1. Patient factors** | **1.1** Denial or minimisation of hearing loss<br>**1.2** Fears and concerns<br>**1.3** Lack of awareness/ knowledge<br>**1.4** Struggles with hearing loss<br>**1.5** Other comorbidities |
| **2. Systemic factors** | **2.1** Patient-clinician relationship<br>**2.2** Clinician knowledge and support<br>**2.3** CI information<br>**2.4** Referral pathway/ health service |
| **3. Social factors** | **3.1** Hearing others' experiences<br>**3.2** Effect of hearing loss on social life<br>**3.3** Support/ encouragement from others<br>**3.4** Ethical issues<br>**3.5** Stigma |

#### Theme 1: Patient factors

##### 1.1 Denial or minimisation of hearing loss

Some participants explained how they minimised the severity of their HL, thinking it was manageable and underestimating how bad it was, so therefore never envisaged they would be eligible for a CI. It was only when they realised the extent of their hearing loss that they had the “drive” to pursue a CI.

> *“I just didn’t think my hearing was in that region, that area where it was so bad to get [a CI]. So I never considered asking about one.”* Participant 3

> *“I think that is why, you know, some people put it off, because they can still manage. And so they haven’t got that drive. It’s when you’ve got nothing to lose- it’s worth it.”* Participant 1

One participant explained how they felt a sense of belonging in the hearing world and were reluctant to let go of this by accepting their HL.

> *“I felt I was part of the hearing world and that was my place and I wasn’t going to let it go.”* Participant 1

##### 1.2 Fears and concerns

Many participants described having fears or concerns about several different aspects of receiving a CI as a reason they were initially hesitant about CIs. One of the fears which came up most commonly was losing residual hearing in the implanted ear. They were worried that in cases where the CI may not work properly, for example issues with the battery, they would be left with no hearing.

> *“I was aware at some point after speaking to [the surgeon] that whichever ear was implanted, then your hearing in that ear would be completely gone. Uh, that was a little bit of a concern. If the implant didn’t work, then I would only have one ear that had some hearing.”* Participant 3

> *“It is such a terrifying thing and, you know, you are one battery away, or one whatever it is, away from losing your hearing completely. The minute you take your processor off at night, it’s gone.”* Participant 1

However, one participant mentioned that when they realised how little hearing was left in their ear anyway, the loss of residual hearing became less of a concern for them.

> *“But the thing that stuck in my mind is… that it can lead to losing hearing in that ear. And at the time I thought, oh, I don’t want to lose my hearing in that ear- the thought of having nothing there. But it soon became apparent that there was so little anyway, if I did then lose the residual hearing, it was not going to be worth keeping.”* Participant 5

Another fear that participants talked about was about the surgery itself, including the anaesthetics, how painful the procedure would be or the recovery afterwards.

> *“I’ve seen some [people] absolutely petrified, you know, sick with concern about having it. If you’ve never been in hospital before, you never had an operation before, it’s an even bigger thing than just having a cochlear implant surgery.”* Participant 5

> *“Everyone thinks… I could have that injection and will never come back to this life. That’s quite a thing to face at any time.”* Participant 2

> *“…in the lead up to the operation, there’s a bit of concern about the operation. How is it going to go? How painful is it going to be? Um, recovery.”* Participant 3

Participants were also concerned about whether the CI would work for them or whether they would be in the small percentage of people for whom the surgery is unsuccessful. This included worries about how much they would be able to hear or understand with the CI.

> *“The thought of the processor not working, the actual cochlear implant I should say, not working right and not functioning. Going to the switch on and them switching it on and they say “can you hear me?” and I just sit there like “well have you switched it on yet?” That for me was the… I wouldn’t say scary thing but that was in the back of my mind, and I wouldn’t say it was a discouragement. It was just me having nagging doubts as to am I doing the right thing.”* Participant 6

> *“I of course was, as I’m sure everybody was or is [concerned about] will it work for me? How much will I actually be able to hear or understand or acquire? Is it actually going to be better?”* Participant 2

One participant mentioned they were concerned that receiving a CI would stop them from being able to try new technologies or treatments for hearing loss in the future.

> *“I was aware of genetic treatments that were underway for the future with hearing loss. Treatments to rejuvenate the hair cells. This kind of thing. And I did wonder if I was being premature, perhaps precluding the use of these at some point in the future.”* Participant 4

##### 1.3 Lack of awareness/knowledge

A lack of knowledge or awareness of CIs by the participants was a common barrier to implantation. Some participants described how they didn’t know what CIs were or that they were an option for people with hearing loss until it was brought up to them by a clinician.

> *“I had not really heard of it before [the consultant mentioned it]… And I certainly wouldn’t have known how to go about [the assessment and referral process].”* Participant 5

> *“That was the first time that I was ever made aware that there was a device other than a hearing aid to help you if you’ve got hearing loss. Up until that point of meeting the consultant, I had no idea about cochlear implants at all.”* Participant 6

There was also a sense of lack of entitlement; another participant who was aware of CIs did not realise they may be eligible for one as they thought it was only available for children or those with a genetic or congenital cause of HL.

> *“I thought it was something for children to give them, you know, a chance of a good life. And for people who were born deaf or had, you know, inherited a deafness from family, or something like that, in their family history. So I didn’t really think I was entitled.”* Participant 3

Participants also spoke about how the vast amount of circulating misinformation could discourage people from receiving a CI.

> *“There’s loads of dreadful myths, and if people don’t get them corrected, they won’t even get as far as getting a referral.”* Participant 1

##### 1.4 Struggles with hearing loss

The participants recounted the many struggles of hearing loss and the effect of hearing loss on their lives as a key motivator for deciding to undergo cochlear implantation. One such struggle was the challenge of managing at work with hearing loss, with participants feeling like they had no other choice but to endure.

> *“[Hearing loss] made work very challenging, very tiring… I was the only person qualified to do a lot of assessments and things so that was one reason why I was able to keep going, because if they didn’t have me, they didn’t have anybody…”* Participant 1

Some participants even had to give up their jobs or retire early due to the effect of hearing loss on their work lives, especially in occupations where it was necessary to communicate regularly with colleagues or clients.

> *“I retired early from work. I retired at 55 because… I was getting to the point where I had to. I had a job where I was quite often in meetings with people, and again it was difficult. A room of maybe six or eight people around the table talking, cross talk across the table. it was getting difficult to keep up with work.”* Participant 3

> *“…as my hearing deteriorated, I couldn’t hear the clients over the phone. And then it got to the point that I couldn’t even hear them properly in a one-on-one room. And…I had to hand my notice in. So that all came to a halt.”* Participant 5

They also reported that using hearing aids were inadequate for their hearing loss.

> *“Even with hearing aids, [keeping up at work] was a problem. I got the hearing aids to try and help with that, but it still wasn’t that great.”* Participant 3

Using the telephone was also a difficult task that many participants couldn’t manage so they often had to rely on others for this.

> *“Hearing loss has affected my ability to use the telephone. And that became quite marked. And it meant that I was pushing the responsibility for making telephone calls to my wife.”* Participant 2

Most participants explained how their relationships with family and friends were negatively affected by hearing loss. For example, one participant explained how hearing loss was causing stress in their relationship with their spouse when they didn’t understand the accommodations that were needed for effective communication.

> *“My hearing loss was affecting my relationship with my wife because despite my saying to her there’s little point in trying to speak from one room to the other because I won’t hear, she would continue to do so… So things started to become… unnecessarily strained.”* Participant 2

Participants also described how there was sometimes a lack of understanding or empathy from relatives and acquaintances about their hearing loss and misconceptions that a hearing aid or cochlear implant is a total cure.

> *“I’ve had people say to me, “You’ve got hearing loss. Well, get on with it,” you know. I tend to avoid people like that anyway, but I think generally people aren’t that perceptive or empathetic when it comes to living with hearing loss.”* Participant 6

> *“With my relationships, I found it increasingly hard to hear my husband… My nephew, niece, who are young at the time, they’ve got high pitched voices, struggling with them. Obviously, friends likewise. So that all had a negative impact because people will turn around to say, well, isn’t your hearing aid working? …So there’s still very much an expectation from friends and family that either a hearing aid or a cochlear implant is the total answer.”* Participant 5

One participant mentioned that their hearing loss was having a negative impact on their mental health due to all the issues that came with it.

> *“There was a period when I was quite depressed because I was trying to adjust to the hearing loss and also the severity of the hearing loss.”* Participant 6

Eventually, participants reached a breaking point where they believed they needed to seek treatment, or they and their families would continue to suffer.

> *“And I thought, either we spend the next 30 years like this or I do something about it …and [my family] are going to suffer as much as me… So, you know, you gotta go for it. Things can’t get any worse really.”* Participant 1

##### 1.5 Other comorbidities

Some participants recounted that their decision to get a CI was delayed by other medical comorbidities they had, which they thought to be more important and so wanted to treat first.

> *“It coincided with having to have further orthopaedic surgery, which was more important at that time. So I delayed having my implant by about a year because of having the other surgery done.”* Participant 5

One participant also described how their comorbidities would make the CI surgery more complex, so this was a concern for the procedure.

> *“I have atrial fibrillation… So there was a bit of a concern about my heart and the anaesthetic and so on. So, you know, I had an ECG before, and the anaesthetist had a look at it and so on. So there was a concern, yes.”* Participant 2

#### Theme 2: Systemic factors

##### 2.1 Patient-clinician relationship

The patient-clinician relationship was an important factor for many of the participants. Having a positive relationship with their audiologist or consultant encouraged many in their decision to undergo implantation and increased confidence in the medical team. Participants placed value in a clinician who was empathetic and worked to build trust with the patient.

> *“…they were always very good, very supportive so that probably did make quite a big difference to me, helping the decision to go ahead and getting the surgery done, the whole bit. It was a positive thing for me.”* Participant 6

> *“His assurance, as it were, and the fact that he was, as I said, such an empathetic person… that was very strong.”* Participant 2

However, on the other hand, some participants talked about experiencing a lack of empathy or support from clinicians, which hindered them in the referral and assessment process. In some cases, there was a feeling of superiority, with the clinician not listening to the patient’s concerns.

> *“Certainly the audiologist I saw needed to listen a bit more to the patient rather than assume that they know better.”* Participant 5

> *“My local GP wasn’t very understanding which is a shame because GPs see a lot of people and a different spread of people and all different illnesses they possibly have, but he didn’t really understand how I was trying to live my life with a hearing aid, with hearing loss, but he was quite… well I wouldn’t say dismissive, but he didn’t really seem interested.”* Participant 6

##### 2.2 Clinician knowledge and support

The knowledge that clinicians had of hearing loss and cochlear implants acted as a motivator or a barrier to participants as there was considerable variability between clinicians. One participant mentioned how the clinicians they first saw about cochlear implants couldn’t advise them at all and they were discouraged from going forward with it. Another spoke about how the clinician initially assessing them did not seem to know the referral criteria well.

> *“They didn’t appear to know… much about them, if anything, and couldn’t advise me. Um, so time went by. Things got somewhat worse. I then think I saw the consultant audiologist at the hospital who said ‘[Participant 2], it’s not appropriate for you and it really wouldn’t work’ - very downbeat.”* Participant 2

> *“I think the complaint is that before I was referred, the person referring me should have checked on the requirements… he didn’t really understand it, he just knew that I was beyond the hearing aids and referred me on that basis.”* Participant 4

Conversely, clinicians who were knowledgeable about cochlear implants and could explain in a clear manner the procedure and technology were greatly appreciated by the participants. In these cases, having understood thoroughly the benefits and risks of cochlear implants, participants felt secure in their decision to go ahead with implantation.

> *“I was really privileged, I believe, to have [Audiologist] as my audiologist, who was prepared when I wanted to ask about how is the electrode stimulated or what is the profile of the voltage change across, between your base, er, level and the implant voltage. And she would get out research papers and send them to me. You know that sort of support was wonderful.”* Participant 2

> *“And [the surgeon] talked me through what would happen, my wife and I, and we were both excited by it. We both thought that this is the thing to do. So we agreed to go ahead… [He] was very good at explaining diagrams and talking us through what would happen.”* Participant 3

##### 2.3 Cochlear implant information

Participants described a strong desire to seek out up-to-date information about cochlear implants prior to, and after, implantation. Aside from clinicians and USAIS, sources included medical textbooks, television, YouTube, journal articles, websites and local/national CI support groups. There was a particular emphasis on finding unbiased and reliable sources.

> *“…looking through the Internet and YouTube cause there’s a lot of stuff on YouTube, which is sort of helpful-ish… I think it’s just finding that information and knowing where to look… you really gotta push yourself and go and knock on the right doors…”* Participant 6

However, one participant was mistrustful of manufacturers’ information and advertising.

> *“I looked at the RNID and Hearing Link websites. I never looked at a manufacturer’s… Deeply suspicious of manufacturers and advertising.”* Participant 1

##### 2.4 Referral pathway/health service

Many participants described challenges they experienced with the referral pathway and healthcare system in general. For example, some participants spoke about a lack of continuity or organisation in two separate areas: firstly, when switching services or hospitals and secondly when being assessed by a different clinician.

> *“I moved hospital and I just fell off the radar.”* Participant 1

> *“[Two different hospitals] were fighting over who should take care of me. So I was kind of abandoned for about six months.”* Participant 4

> *“And every time you went, you saw a different audiologist- there was no continuity. So it wasn’t a great experience.”* Participant 5

Another point which came up frequently was about the state of the NHS in general and how problems with staffing or funding meant operations were cancelled or pushed back, which led to further delays in getting implanted.

> *“Bed got cancelled a couple of times. Usual thing on the NHS… but unfortunately, we know that’s commonplace… I think it’s just a matter of funding, staffing and so on.”* Participant 5

> *“I was gonna have it done at [hospital] but they cancelled on me because the microscope that they use in surgery, apparently it was broken or damaged, it wasn’t functioning, so they cancelled on me once and the second time, I think there was an issue with some staff.”* Participant 6

In terms of referral pathway, a systemic lack of knowledge about implants meant that participants were not made aware that implants were an option for hearing loss or referred on for further assessment. One participant emphasised the need for better training for referrers about the referral criteria.

> *“…had it not been for my enquiring mind, I would have found [being referred] pretty difficult. Because the audiology department was not, I would have said, sort of positive to the idea and possibly the routine audiology staff were not aware of them at the time.”* Participant 2

> *“I suspect when they change the requirements, it needs to be fed back to the referring people, you know, the referring audiologists. So to that extent, I think there’s a need for better communication.”* Participant 4

However, aside from the initial troubles with getting referred, participants found that the rest of the process ran quickly and smoothly, although there was a comment from one participant that the hearing tests were a bit stressful.

> *“I found it incredibly quick… Within two weeks of me seeing the consultant I’d got the pack from Southampton and the first set of appointments through…”* Participant 1

> *“Everything seemed to go as they told me it would go. The different stages, it was quite easy… it’s quite stressful because you’re doing hearing tests, and you can’t hear, you know? So you’re listening to tiny little beeps and half imagining them. So it’s a little bit stressful, but that’s not because of the process. That’s just because of what you’re trying to do. So yeah, little bit stressful but no, it was very very straightforward and the process was quite easy.”* Participant 3

The participants praised the staff and service at USAIS, commenting that the support throughout the process, especially the expertise and friendliness of staff, was excellent.

> *“Everyone at USAIS, including consultants to all the audiologists etcetera, you cannot fault. So they’re just all amazing. And all the support I got… everyone couldn’t do enough to help and reassure and make sure that we were doing the right thing.”* Participant 5

#### Theme 3: Social factors

##### 3.1 Hearing others’ experiences

One of the major motivators to implantation reported by most of the participants was speaking to others with an implant and hearing their experiences. Seeing and hearing stories of success made the participants feel more reassured about receiving an implant themselves.

> *“…having read about it online and looked at other people’s experiences online, cochlear website… how patients talk about it… And I was very impressed with how for most people it was very, very successful. And, you know, I wanted that. I thought, well, this, this could be amazing. So that was… that was definitely what motivated me, other people’s experiences and the general idea that it was usually pretty successful.”* Participant 3

> *“I talked to people there who had had their implant, and obviously their experience was very positive. And that led me then to think, OK, you know, this is something I’m going to pursue. But certainly, talking to people who already had that and had been on that journey was a major deciding factor.”* Participant 5

One participant even suggested that sending a short video of people with cochlear implants to potential cochlear implant candidates early in the assessment process could be a source of encouragement for them to pursue an implant.

> *“Southampton has a set of little films you could look at from people with [a CI]. And I think if that was sent along to you instantly, you know, as the first thing you got, ‘we’ll be sending your information pack soon, but here’s some films from people who’ve had them’ might possibly even be a bit better.”* Participant 1

##### 3.2 Effect of hearing loss on social life

The difficulties of enduring hearing loss in their social lives were also a driver to implantation. Many participants reported how they felt socially isolated from family and friends and how challenging it was to feel included in social situations due to the major problems in communication they were experiencing. This also included being unable to do their usual hobbies, such as attending the theatre.

> *“[During] a family meal, I would generally be left out… Not deliberately, but everybody’s talking. You’re probably aware of it, but anybody who’s deaf, it’s very difficult to follow one conversation, never mind two or three. Um, if you lip read, you can look at somebody who’s talking, but by the time you turn to the person who’s answering to lipread them, they’re finished or they’re halfway through it. So it’s very difficult to follow those conversations so I suppose there’s a loneliness comes from that. You feel a bit isolated, a bit on your own.”* Participant 3

> *“…my social world involves around my friends, family, theatre, you know, and that sort of thing. And I felt that as my hearing deteriorated my world just shrunk further. Um, so there was even more and more things that I then couldn’t do. And to sit on the edge of a conversation with friends or family in a proper restaurant and just not know what’s being said, by whom, and you feel just like a bystander watching on.”* Participant 5

These feelings of loneliness and isolation and a desire to be involved in society again were key deciding factors for people in seeking treatment for their hearing loss.

> *“I wanted to engage with people again because I was born a hearing person… And so, it was just being able to engage with people and connect with people regularly so that I didn’t feel isolated and cut off… that was my primary reason [to seek treatment for hearing loss].”* Participant 6

##### 3.3 Support/encouragement from others

Often, the push to get a hearing test or to seek treatment came from family or friends. Having a strong support network also made the implantation and recovery process a lot easier for participants.

> *“One of our good friends is a very positive lady who doesn’t let things lie if they need to be addressed. And she said [Participant 2], your hearing is getting in the way of social events. And look, Doctor X has a clinic just down the road… you ought to go and I’m going to make you an appointment.’”* Participant 2

> *“I had plenty of support at home and encouraged to do my daily rehab exercises and my husband went with me to most of my appointments or my father did, came to a couple as well. So, completely supported by family and friends”* Participant 5

##### 3.4 Ethical issues

A few ethical problems were brought up in the interviews, most revolving around issues with cochlear implantation in the Deaf community. Participants commented on the pressure on members of the Deaf community to forgo implantation as it is seen as taking away from their identity as a Deaf person.

> *“I think there’s a lot of peer pressure on most of the Deaf not to have a cochlear implant because implanted people leave the Deaf community, so they see their community shrinking all the time.”* Participant 4

> *“I know a lot of people who decided not to go ahead with it, one or two friends that I know in the Deaf community, there’s a lot of… it’s not resentment but they feel threatened that they are somehow going to lose their identity because they have a CI.”* Participant 6

##### 3.5 Stigma

A minor barrier that some participants mentioned was the stigma of the appearance of a CI and being self-conscious about “funny looks” they would get from people. Another participant described how even manufacturing companies perpetuate the stigma by emphasising the need to hide a hearing aid or CI.

> *“The negative thing was I found when I first got my CI, the processor, I used to have an old Cochlear, and it used to have a coil thing and the whole bit… I used to get a lot of people giving me funny looks and then they sort of look away, say something to a friend and then they do that again… I was very, very conscious when I got my first processor. Just how much this thing was sat on the side of my head because you have the whole cable, and you used to have the ear thing to fit over your ear. I found that quite cumbersome.”* Participant 6

> *“…this whole stigma about hiding, and I think the adverts for hearing aids are particularly bad at it- we have this amazing hearing aid, no one will see it. Well, they just create the stigma.”* Participant 5

## DISCUSSION

Multiple factors contribute to the low uptake of cochlear implants for adults in the UK (9). This study highlights the key barriers and motivators that patients, over the age of 60 at USAIS, experienced during their journey to being implanted. Key barriers included socioeconomic status, lack of knowledge about cochlear implants amongst both patients and clinicians, problems in the referral process, personal denial or minimisation of their hearing loss and fear of surgery or the implant not working. We found most individuals who underwent implantation at USAIS were from higher socioeconomic groups despite the burden of hearing loss being greater in lower socioeconomic groups (28). This suggests that being of lower socioeconomic status may be a barrier to implantation. Certain services and areas referred more patients than others suggesting a geographical effect. Support from family, friends and clinicians, peer support and the severity of the impact of hearing loss on daily life were identified as key drivers to proceeding with cochlear implantation.

This is the first study to investigate barriers to cochlear implant uptake at USAIS. The need to better understand the barriers to cochlear implant uptake in older adults was highlighted by involvement with our PPIE group which led to the design and implementation of this study. A strength of this work was the mixed-methods approach which included a service evaluation to evaluate the demographical profile and routes to referral for patients at USAIS and semi-structured interviews to gather a detailed perspective of people with lived experiences of the process. We collected rich qualitative data which is beneficial in identifying what might improve health outcomes by enabling more eligible people to undergo cochlear implantation at USAIS (29). A limitation of this study is that all study participants went on to receive a cochlear implant therefore we have not included data from people who may have been referred to the service and found to meet the eligibility criteria, but who did not go forward with implantation. The perceived barriers for proceeding with implantation from those who did not proceed at USAIS are therefore not known. We also did not include people who may be eligible, according to NICE candidacy criteria, for a cochlear implant but who had not been referred to the service. Our analysis demonstrates that geographical disparities influence how patients are referred to the service. However, further investigation is required to understand the breadth of barriers to referral to USAIS.

Our study adds to the evidence that socioeconomic status influences cochlear implant uptake. Previous studies have reported that attendance of initial cochlear implant candidacy assessment appointments (30) and uptake of implants (31,32) is affected by socioeconomic status. This may be due to more pressured healthcare services in deprived areas. In addition, the financial costs of implantation borne by the individual, such as travel costs to attend appointments, time off work, and loss of earnings may have an impact (10). Our findings demonstrated that sex and ethnicity were not key barriers to cochlear implant uptake in the patient cohort at USAIS. However, other studies in the UK found that older males and patients from Asian or black ethnic background were less likely to be referred for cochlear implantation (13). The limited diversity in the USAIS catchment area limits our ability to comment on ethnicity as a barrier. Despite what we have found at USAIS, increasing the ethnic diversity of CI recipients across the UK should be an area of focus. We found differences in referral rates between services and areas suggesting that both the area you live in and the healthcare professional you see, may act as a barrier or enabler to cochlear implant uptake.

In the UK, there are audiologists based in secondary care units who are designated ‘cochlear implant champions’. These individuals receive additional training in cochlear implants and raise awareness and support referral to CI services (33). The presence of effective champions within Dorset and Hampshire is a likely contributor to the disproportionately high referral rates from these areas. Additionally, proximity to the CI service may be a contributing factor. The ability, or inability, to travel to regional implant centres has been described as a barrier, particularly for older adult patients (34). Similarly, poorer knowledge of cochlear implants and candidacy criteria in certain areas could explain lower referral rates rates (9,35–38). A finding from our qualitative analyses that a knowledgeable clinician and a good patient-clinician relationship was a motivator supports existing findings (10,39).

Spending more time counselling patients about the success and failure rates of implantation and the low risk of complications may help them overcome their hesitancy (40). Key motivators to taking up an implant in our study were the desire to try something to overcome the daily struggles associated with hearing loss and having support from clinicians, family and friends and other people with implants. Knowing someone with an implant and having the opportunity to learn from their experiences has been described previously as a motivator to uptake (14). This highlights the value of advocacy that is driven by people with lived experience (41,42).

This research could inform the development and implementation of strategies to improve national access to, and uptake of, cochlear implants for older adults. At USAIS, we now use community engagement as a strategy to raise awareness of CIs in our local area area (43). Researchers have been visiting community groups including a hearing loss support group and groups for underserved communities, to get to know members, build trust and share and discuss information about hearing health and cochlear implantation. This approach has led to people from a diversity of backgrounds to join our PPIE group. This has created a conduit for new knowledge between communities and researchers. A similar approach could provide more support to local clinicians in assessing and referring patients who may be eligible for cochlear implants. Evidence from this and other studies suggest that socioeconomic status is a major factor in referrals to services and uptake of cochlear implants. At a local level, more financial help could be offered to people from disadvantaged backgrounds. At a national level, healthcare policies and resource allocation need to be reviewed to resolve health disparities between different geographical regions. Further studies to identify persisting barriers will support this effort (44).

There are several unanswered questions that require further research. This includes 1). Why are there disparities in referrals between districts in the service catchment area and what might reduce these disparities? 2). What are the barriers for people who are referred to the service and meet implantation criteria but choose not to have an implant? 3). Does community engagement improve, or diversify, who gets access to cochlear implantation? 4). What are local clinicians’ understanding and views of cochlear implants and their role in assisting patients access them? How can we support them in their discussions with patients about cochlear implants and improve their knowledge about referral criteria?

## Data Availability

Datasets from this study will be published in the University of Southampton PURE repository.

## Acknowledgements

The authors are grateful to members of ALL_EARS@UoS patient and public involvement and engagement (PPIE) group who continue to give up their time to contribute to the group. Thank you for contributing to discussions in meetings and for sharing your lived experience of deafness and hearing loss to support and shape our research.

## Data Statement

Datasets from this study will be published in the University of Southampton PURE repository.

## Author Contributions

All authors contributed to conceptualisation and methodology. AG: Investigation, data curation and visualisation, formal analysis, writing – original draft. KH, CF, MG, TAN: writing – review and editing. KH, MG, TAN: Supervision. TAN: Validation. All authors approved the final version. TAN is the guarantor.

