## Supplementary files for "Identification of the motivators and barriers to cochlear implantation in adults over 60 years at an auditory implant centre in the UK: a mixed-methods study"

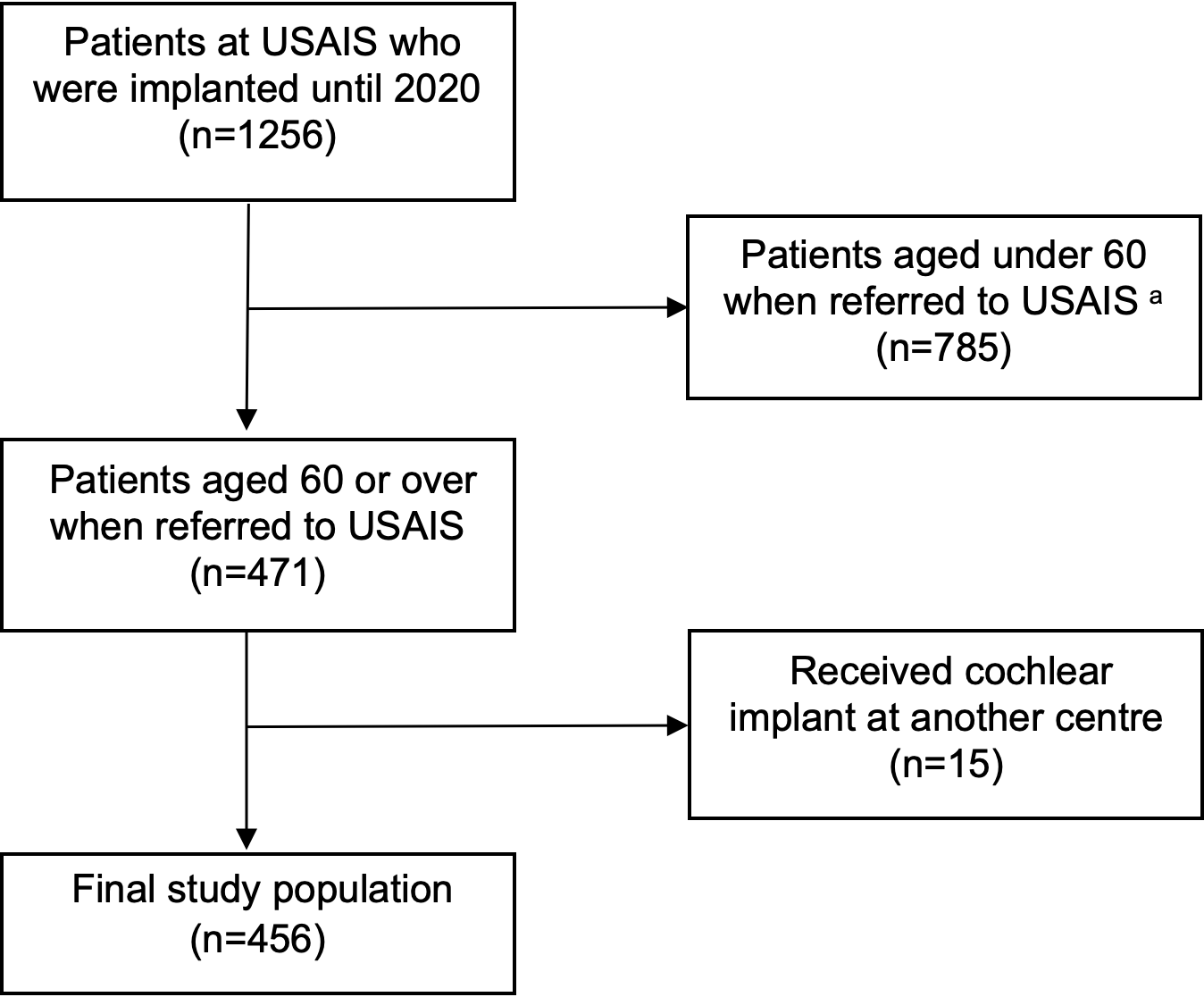

**Supplementary figure 1**. Diagram detailing inclusion and exclusion criteria for the study population.

^a^ Where age at referral was unknown, patient’s age at implantation was used instead to include or exclude them.

**The STROBE reporting checklist**

Title of paper: **Identification of the motivators and barriers to cochlear implantation in adults over 60 years at an auditory implant centre in the UK: a mixed-methods study**

|  | Item Description | Location (or reason for not reporting) |
| --- | --- | --- |
| **Title and abstract** |  |  |
| [1a. Indicate the study’s design](https:/resources.equator-network.org/reporting-guidelines/strobe/items/title-abstract-indicate-study-design.html?utm_source=strobe&utm_medium=checklist&utm_campaign=1_1) | Indicate the study’s design with a commonly used term in the title or the abstract. | Title and abstract |
| [1b. Abstract](https:/resources.equator-network.org/reporting-guidelines/strobe/items/abstract.html?utm_source=strobe&utm_medium=checklist&utm_campaign=1_1) | Provide in the abstract an informative and balanced summary of what was done and what was found. | Abstract |
| **Introduction** |  |  |
| [2. Background / rationale](https:/resources.equator-network.org/reporting-guidelines/strobe/items/background-rationale.html?utm_source=strobe&utm_medium=checklist&utm_campaign=1_1) | Explain the scientific background and rationale for the investigation being reported. | Introduction |
| [3. Objectives](https:/resources.equator-network.org/reporting-guidelines/strobe/items/objectives.html?utm_source=strobe&utm_medium=checklist&utm_campaign=1_1) | State specific objectives, including any prespecified hypotheses. | Introduction |
| **Methods** |  |  |
| [4. Study design](https:/resources.equator-network.org/reporting-guidelines/strobe/items/study-design.html?utm_source=strobe&utm_medium=checklist&utm_campaign=1_1) | Present key elements of study design early in the paper. | Abstract, article summary |
| [5. Setting](https:/resources.equator-network.org/reporting-guidelines/strobe/items/setting.html?utm_source=strobe&utm_medium=checklist&utm_campaign=1_1) | Describe the setting, locations, and relevant dates, including periods of recruitment, exposure, follow-up, and data collection. | Methods – service evaluation |
| [6a. Eligibility criteria](https:/resources.equator-network.org/reporting-guidelines/strobe/items/eligibility-criteria.html?utm_source=strobe&utm_medium=checklist&utm_campaign=1_1) | **Cohort study:** Give the eligibility criteria, and the sources and methods of selection of participants. Describe methods of follow-up. **Case-control study:** Give the eligibility criteria, and the sources and methods of case ascertainment and control selection. Give the rationale for the choice of cases and controls. **Cross-sectional study:** Give the eligibility criteria, and the sources and methods of selection of participants. | Methods – service evaluation |
| [6b. Matching criteria](https:/resources.equator-network.org/reporting-guidelines/strobe/items/matching-criteria.html?utm_source=strobe&utm_medium=checklist&utm_campaign=1_1) | **Cohort study:** For matched studies, give matching criteria and number of exposed and unexposed. **Case-control study:** For matched studies, give matching criteria and the number of controls per case. | N/A |
| [7. Variables](https:/resources.equator-network.org/reporting-guidelines/strobe/items/variables.html?utm_source=strobe&utm_medium=checklist&utm_campaign=1_1) | Clearly define all outcomes, exposures, predictors, potential confounders, and effect modifiers. Give diagnostic criteria, if applicable. | Methods – service evaluation |
| [8. Data sources / measurement](https:/resources.equator-network.org/reporting-guidelines/strobe/items/data-sources-measurement.html?utm_source=strobe&utm_medium=checklist&utm_campaign=1_1) | For each variable of interest give sources of data and details of methods of assessment (measurement). Describe comparability of assessment methods if there is more than one group. | Methods – service evaluation |
| [9. Bias](https:/resources.equator-network.org/reporting-guidelines/strobe/items/bias.html?utm_source=strobe&utm_medium=checklist&utm_campaign=1_1) | Describe any efforts to address potential sources of bias. | Methods – service evaluation |
| [10. Study size](https:/resources.equator-network.org/reporting-guidelines/strobe/items/study-size.html?utm_source=strobe&utm_medium=checklist&utm_campaign=1_1) | Explain how the study size was arrived at. | Methods – service evaluation |
| [11. Quantitative variables](https:/resources.equator-network.org/reporting-guidelines/strobe/items/quantitative-variables.html?utm_source=strobe&utm_medium=checklist&utm_campaign=1_1) | Explain how quantitative variables were handled in the analyses. If applicable, describe which groupings were chosen, and why. | Methods – service evaluation |
| [12a. Statistical methods](https:/resources.equator-network.org/reporting-guidelines/strobe/items/statistical-methods-description.html?utm_source=strobe&utm_medium=checklist&utm_campaign=1_1) | Describe all statistical methods, including those used to control for confounding. | Methods – service evaluation |
| [12b. Statistical methods – subgroups and interactions](https:/resources.equator-network.org/reporting-guidelines/strobe/items/statistical-methods-subgroups-interactions.html?utm_source=strobe&utm_medium=checklist&utm_campaign=1_1) | Describe any methods used to examine subgroups and interactions. | Methods – service evaluation |
| [12c. Statistical methods – missing data](https:/resources.equator-network.org/reporting-guidelines/strobe/items/statistical-methods-missing-data.html?utm_source=strobe&utm_medium=checklist&utm_campaign=1_1) | Explain how missing data were addressed. | Figure 2 legend. |
| [12di. Statistical methods – loss to follow-up](https:/resources.equator-network.org/reporting-guidelines/strobe/items/statistical-methods-loss-to-follow-up.html?utm_source=strobe&utm_medium=checklist&utm_campaign=1_1) | **Cohort study:** If applicable, describe how loss to follow-up was addressed. | N/A |
| [12dii. Statistical methods – matching cases and controls](https:/resources.equator-network.org/reporting-guidelines/strobe/items/statistical-methods-matching-cases-controls.html?utm_source=strobe&utm_medium=checklist&utm_campaign=1_1) | **Case-control study:** If applicable, explain how matching of cases and controls was addressed. | N/A |
| [12diii. Statistical methods – sampling strategy](https:/resources.equator-network.org/reporting-guidelines/strobe/items/statistical-methods-analytical-methods-sampling-strategy.html?utm_source=strobe&utm_medium=checklist&utm_campaign=1_1) | **Cross-sectional study:** If applicable, describe analytical methods taking account of sampling strategy. | N/A |
| [12e. Statistical methods – sensitivity analyses](https:/resources.equator-network.org/reporting-guidelines/strobe/items/statistical-methods-sensitivity-analyses.html?utm_source=strobe&utm_medium=checklist&utm_campaign=1_1) | Describe any sensitivity analyses. | N/A |
| **Results** |  |  |
| [13a. Participant numbers](https:/resources.equator-network.org/reporting-guidelines/strobe/items/participants-numbers.html?utm_source=strobe&utm_medium=checklist&utm_campaign=1_1) | Report the numbers of individuals at each stage of the study—e.g., numbers potentially eligible, examined for eligibility, confirmed eligible, included in the study, completing follow-up, and analysed; Consider use of a flow diagram. | Supplementary figure 1 |
| [13b. Participants – non-participation](https:/resources.equator-network.org/reporting-guidelines/strobe/items/participants-non-participation.html?utm_source=strobe&utm_medium=checklist&utm_campaign=1_1) | Give reasons for non-participation at each stage. | Supplementary figure 1 |
| [13c. Participants – flow diagram](https:/resources.equator-network.org/reporting-guidelines/strobe/items/participants-flow-diagram.html?utm_source=strobe&utm_medium=checklist&utm_campaign=1_1) | Consider use of a flow diagram. | Supplementary figure 1 |
| [14a. Descriptive data – participant characteristics](https:/resources.equator-network.org/reporting-guidelines/strobe/items/descriptive-data-participant-characteristics.html?utm_source=strobe&utm_medium=checklist&utm_campaign=1_1) | Give characteristics of study participants (e.g., demographic, clinical, social) and information on exposures and potential confounders. Present the information in a table. | Results – service evaluation, table 1 |
| [14b. Descriptive data – missing data](https:/resources.equator-network.org/reporting-guidelines/strobe/items/descriptive-data-missing-data.html?utm_source=strobe&utm_medium=checklist&utm_campaign=1_1) | Indicate the number of participants with missing data for each variable of interest. | N/A |
| [14c. Descriptive data – follow-up time](https:/resources.equator-network.org/reporting-guidelines/strobe/items/descriptive-data-follow-up-time.html?utm_source=strobe&utm_medium=checklist&utm_campaign=1_1) | **Cohort study:** Summarise follow-up time—e.g., average and total amount. | N/A |
| [15. Outcome data](https:/resources.equator-network.org/reporting-guidelines/strobe/items/outcome-data.html?utm_source=strobe&utm_medium=checklist&utm_campaign=1_1) | **Cohort study:** Report numbers of outcome events or summary measures over time. **Case-control study:** Report numbers in each exposure category, or summary measures of exposure. **Cross-sectional study:** Report numbers of outcome events or summary measures. | Results – service evaluation |
| [16a. Main results](https:/resources.equator-network.org/reporting-guidelines/strobe/items/main-results.html?utm_source=strobe&utm_medium=checklist&utm_campaign=1_1) | Give unadjusted estimates and, if applicable, confounder-adjusted estimates and their precision (e.g., 95% confidence intervals). Make clear which confounders were adjusted for and why they were included. | Results – service evaluation |
| [16b. Main results – category boundaries](https:/resources.equator-network.org/reporting-guidelines/strobe/items/main-results-category-boundaries.html?utm_source=strobe&utm_medium=checklist&utm_campaign=1_1) | Report category boundaries when continuous variables were categorised. | Results – service evaluation table 1 |
| [16c. Main results – risk](https:/resources.equator-network.org/reporting-guidelines/strobe/items/main-results-risk.html?utm_source=strobe&utm_medium=checklist&utm_campaign=1_1) | If relevant, consider translating estimates of relative risk into absolute risk for a meaningful time period. | N/A |
| [17. Other analyses](https:/resources.equator-network.org/reporting-guidelines/strobe/items/other-analyses.html?utm_source=strobe&utm_medium=checklist&utm_campaign=1_1) | Report other analyses done—e.g., analyses of subgroups and interactions, and sensitivity analyses. | N/A |
| **Discussion** |  |  |
| [18. Key results](https:/resources.equator-network.org/reporting-guidelines/strobe/items/key-results.html?utm_source=strobe&utm_medium=checklist&utm_campaign=1_1) | Summarise key results with reference to study objectives. | Discussion |
| [19. Limitations](https:/resources.equator-network.org/reporting-guidelines/strobe/items/limitations.html?utm_source=strobe&utm_medium=checklist&utm_campaign=1_1) | Discuss limitations of the study, taking into account sources of potential bias or imprecision. Discuss both direction and magnitude of any potential bias. | Article summary  Discussion – paragraph 2. |
| [20. Interpretation](https:/resources.equator-network.org/reporting-guidelines/strobe/items/interpretation.html?utm_source=strobe&utm_medium=checklist&utm_campaign=1_1) | Give a cautious overall interpretation considering objectives, limitations, multiplicity of analyses, results from similar studies, and other relevant evidence. | Discussion |
| [21. Generalisability](https:/resources.equator-network.org/reporting-guidelines/strobe/items/generalisability.html?utm_source=strobe&utm_medium=checklist&utm_campaign=1_1) | Discuss the generalisability (external validity) of the study results. | Discussion |
| **Other information** |  |  |
| [22. Funding](https:/resources.equator-network.org/reporting-guidelines/strobe/items/funding.html?utm_source=strobe&utm_medium=checklist&utm_campaign=1_1) | Give the source of funding and the role of the funders for the present study and, if applicable, for the original study on which the present article is based. | Funding section |

**Supplementary figure 2.** The STROBE reporting checklist.

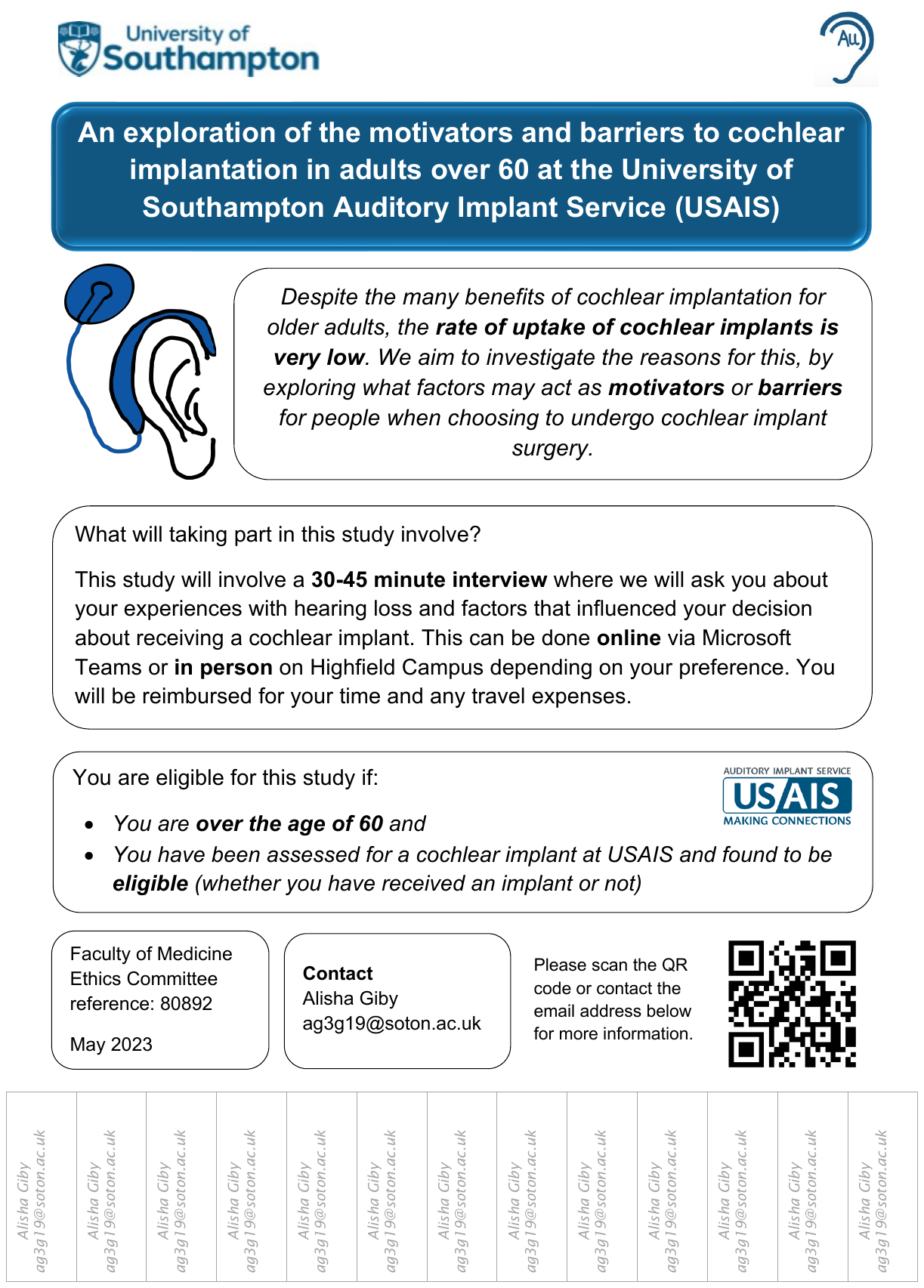

**Supplementary figure 3.** Poster advertising the research study.

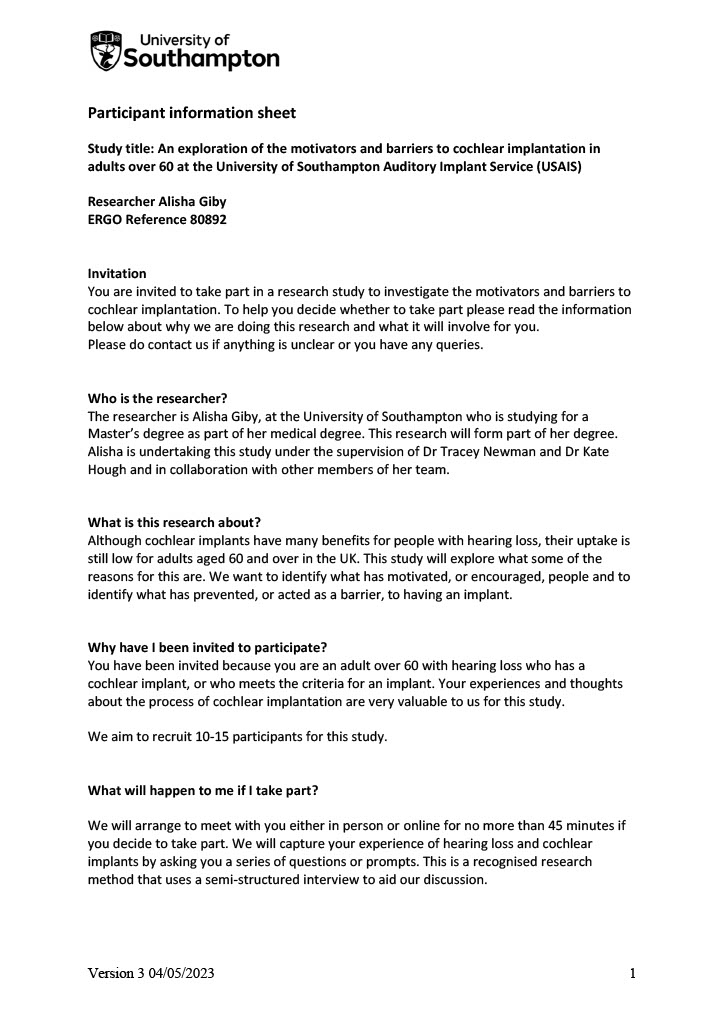

### Appendix D – Participant Information Sheet

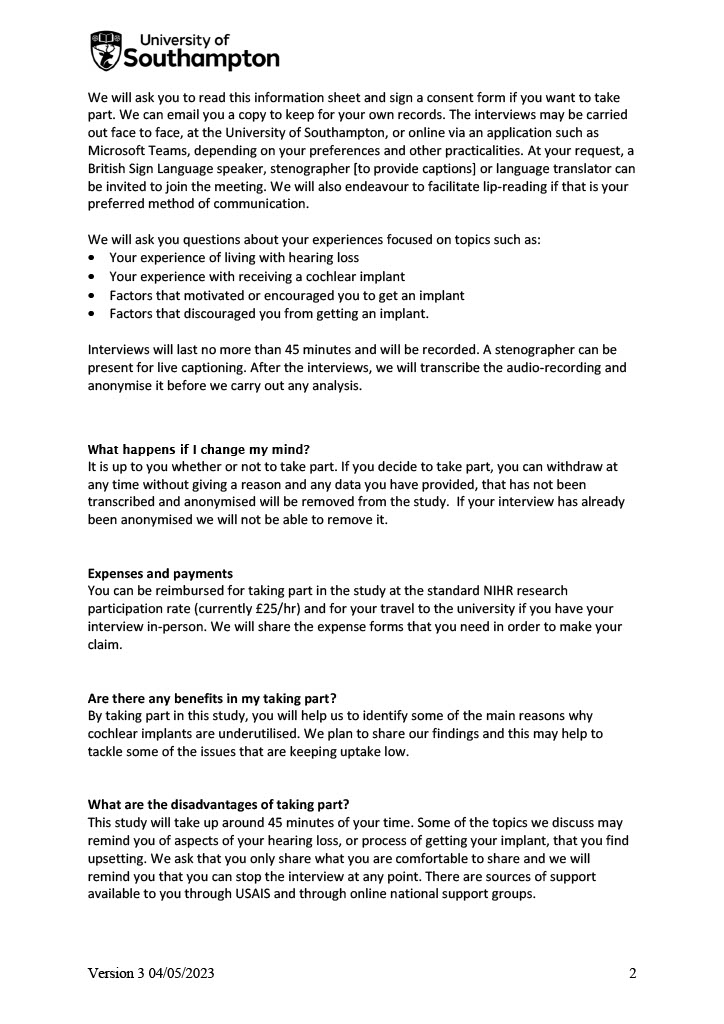

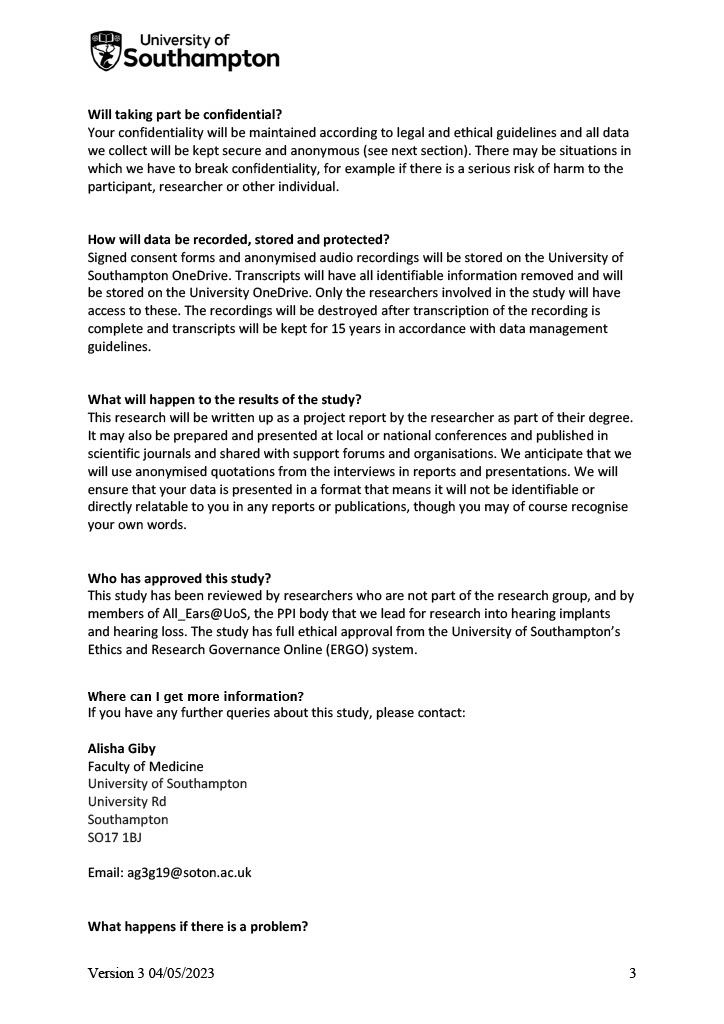

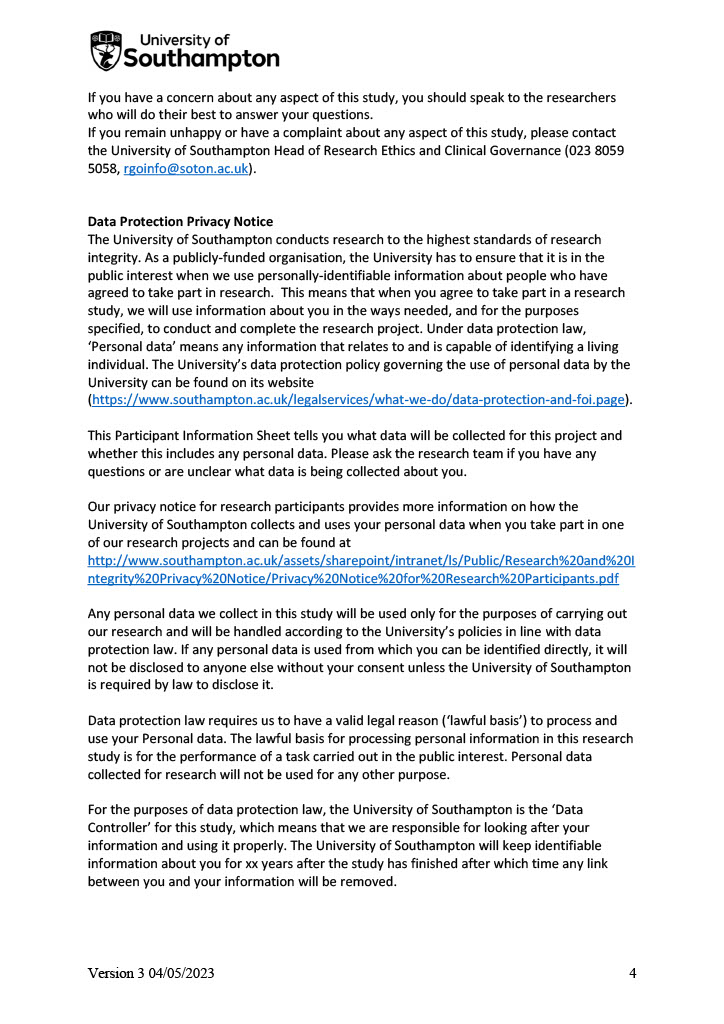

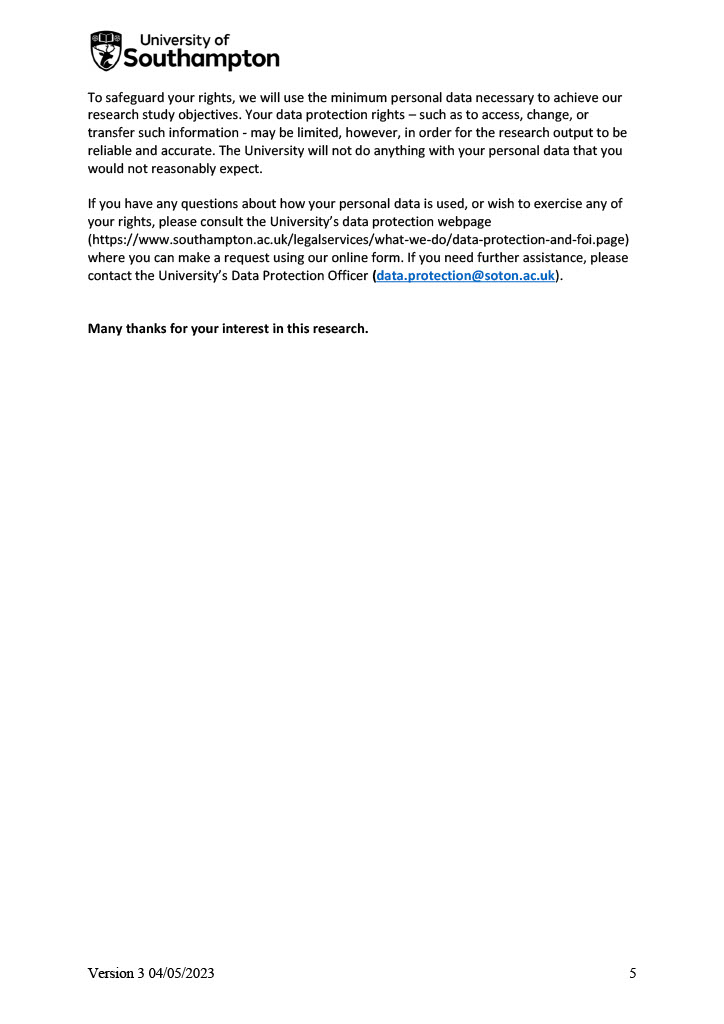
**Supplementary figure 4**. Participant information sheet for participants of the interviews.

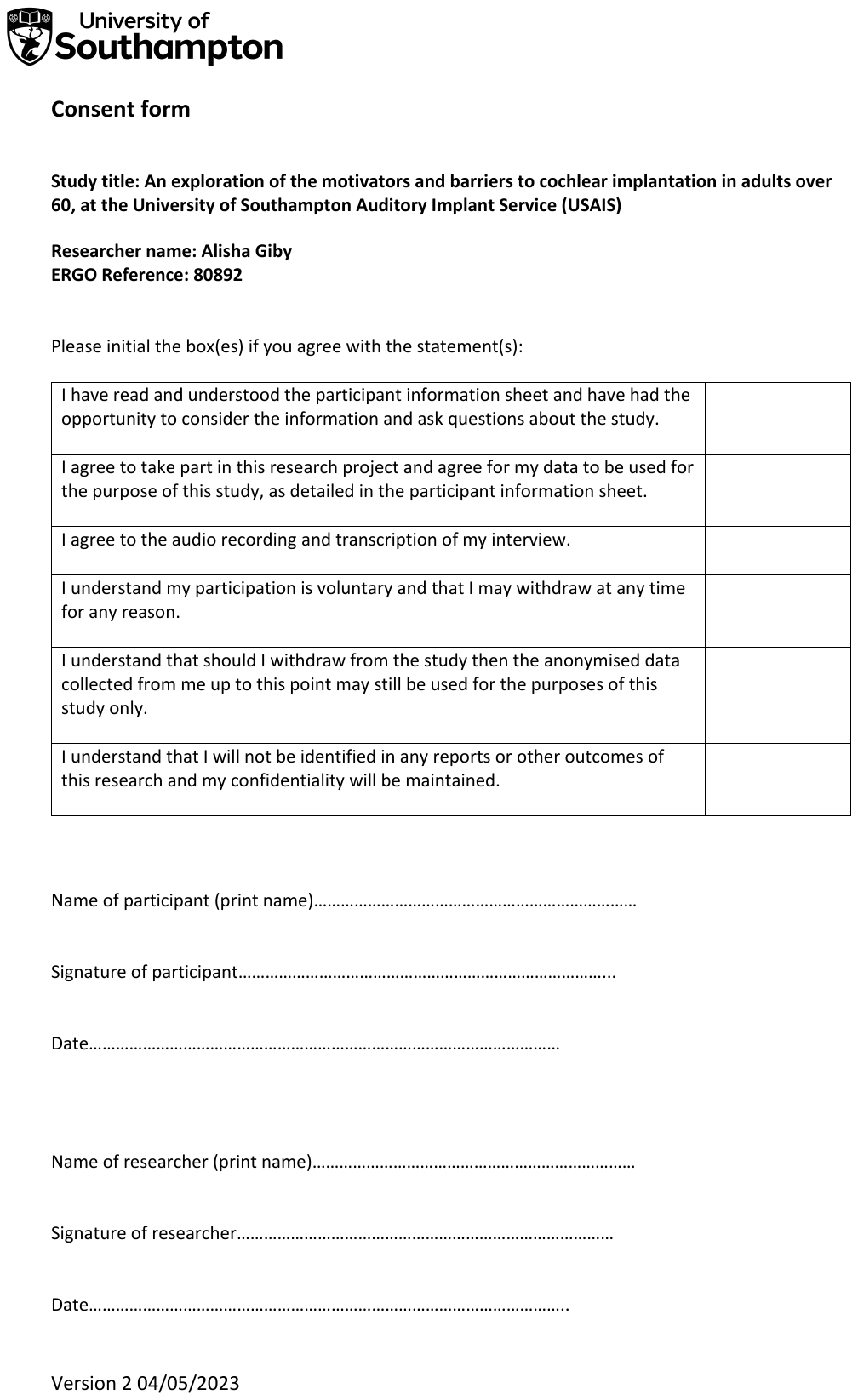

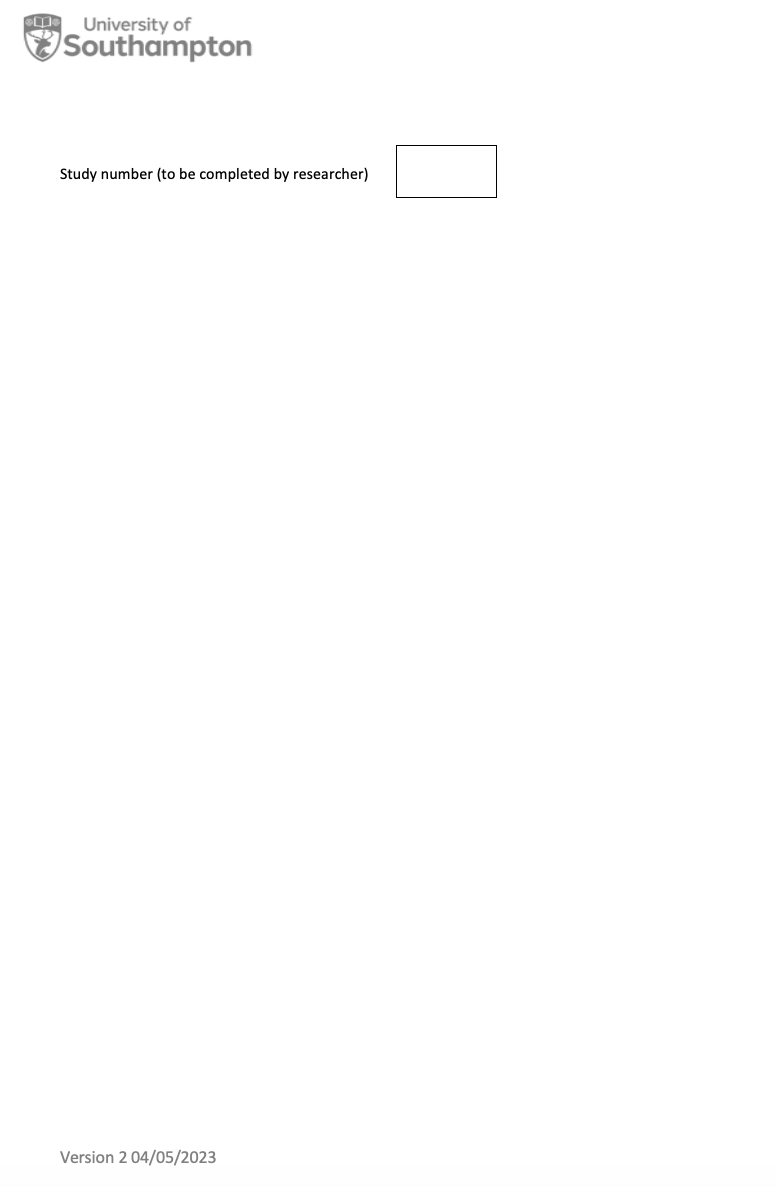

**Supplementary figure 5.** Consent form for participants of the interviews.

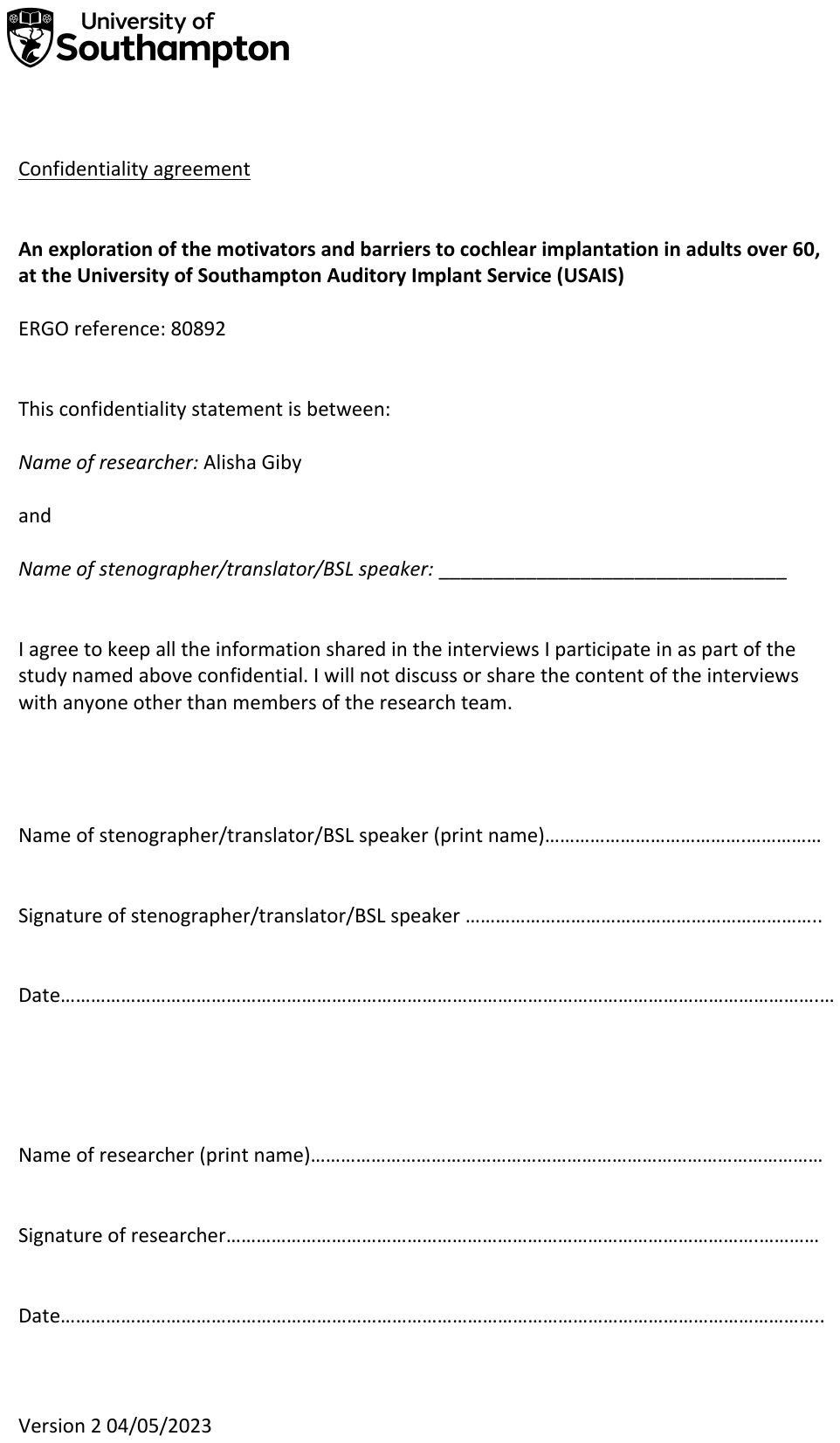

**Supplementary figure 6.** Confidentiality agreement for participants of the interviews.

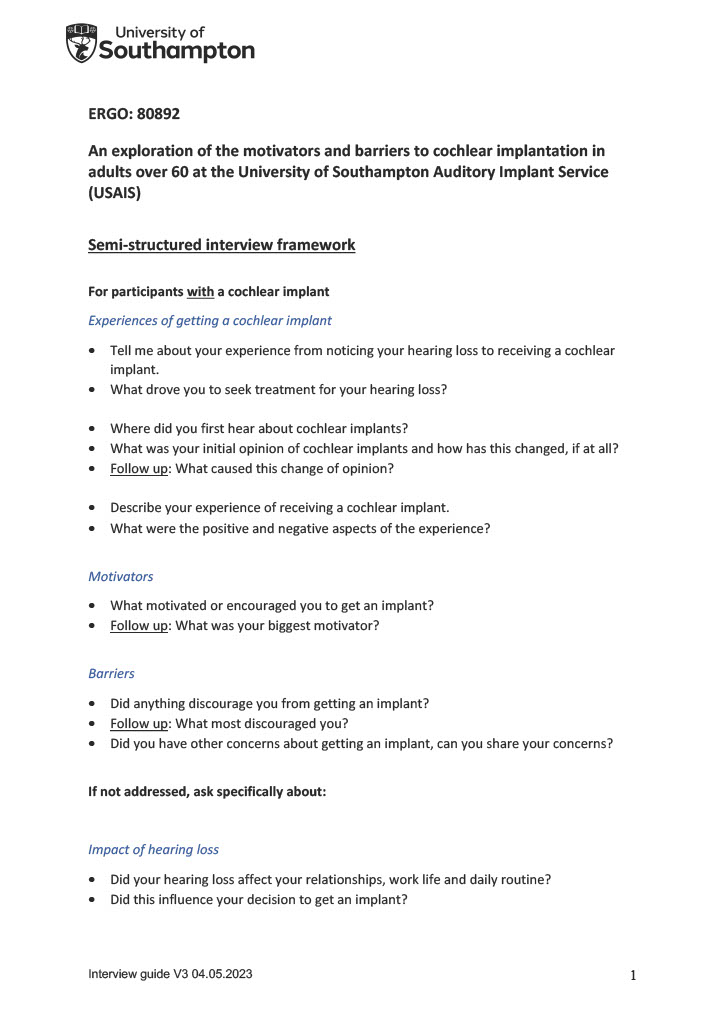

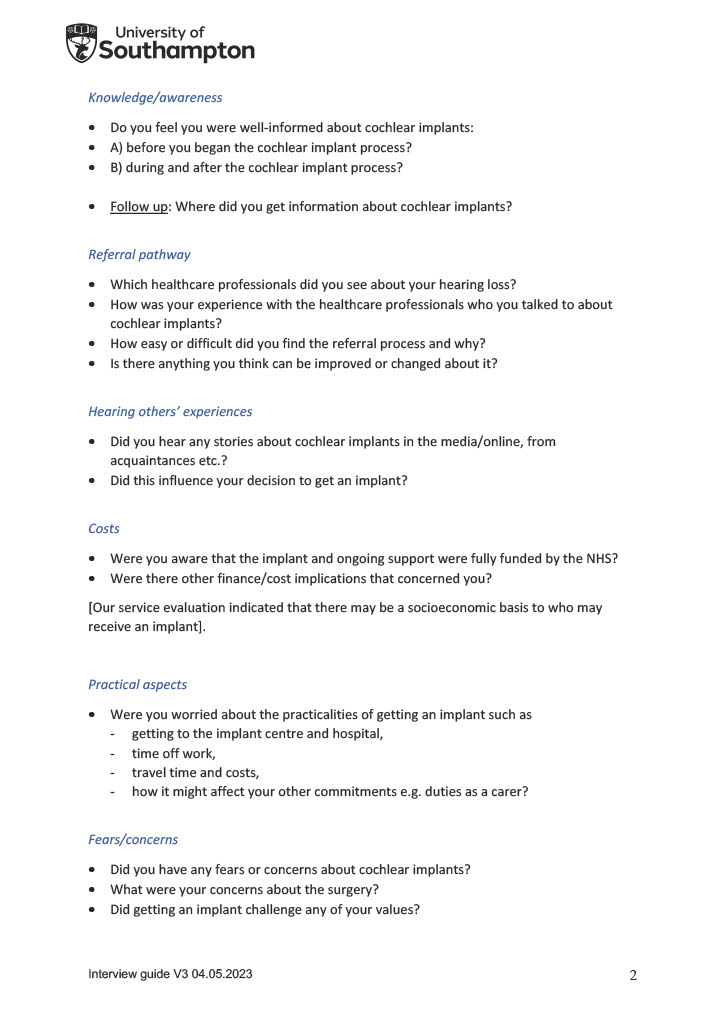

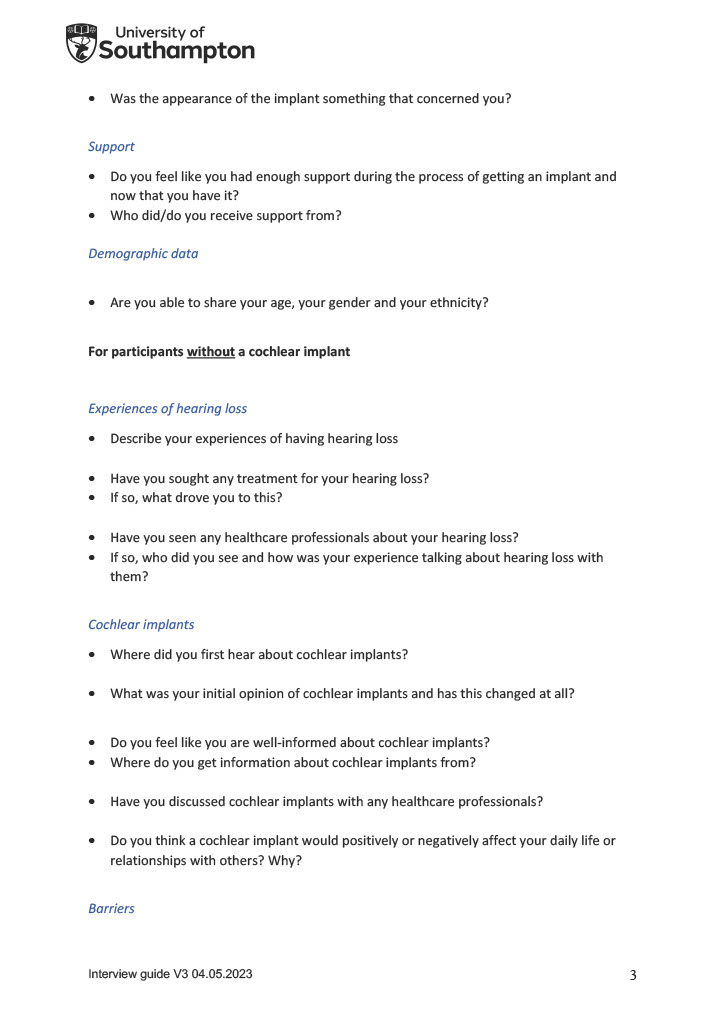

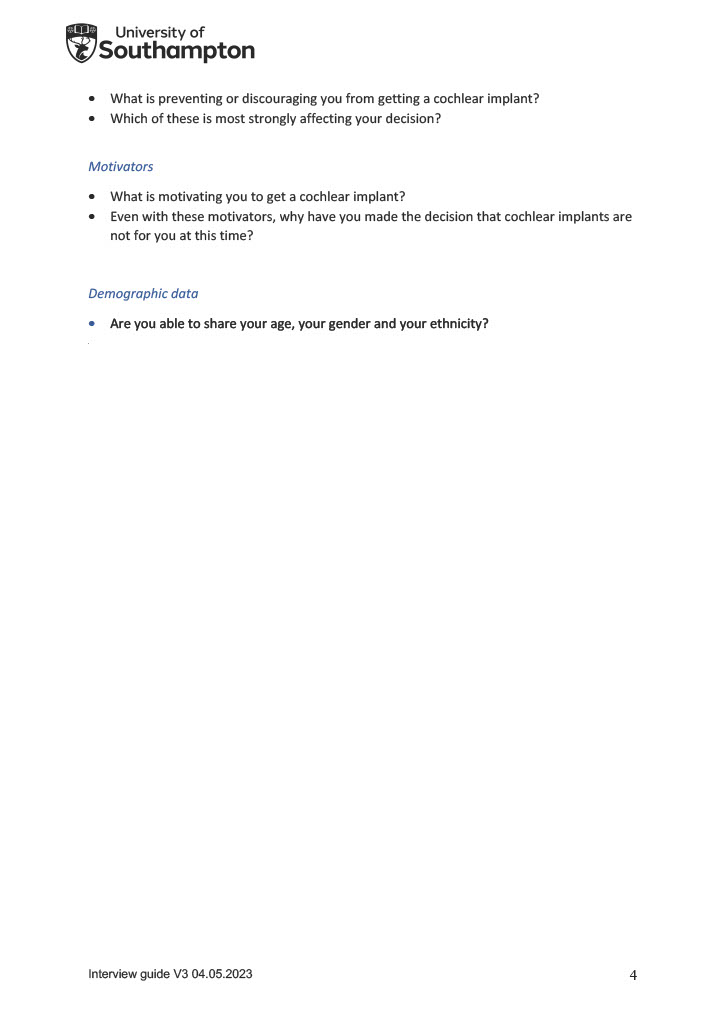
**Supplementary figure 7.** Semi-structured interview guide
